# Effect of storage conditions on SARS-CoV-2 RNA quantification in wastewater solids

**DOI:** 10.1101/2021.05.04.21256611

**Authors:** Adrian Simpson, Aaron Topol, Bradley White, Marlene K. Wolfe, Krista Wigginton, Alexandria B. Boehm

**Affiliations:** Verily Life Sciences, South San Francisco, 94080, California, United States; Civil and Environmental Engineering, Stanford University, Stanford, California, United States; Civil and Environmental Engineering, University of Michigan, Ann Arbor, Michigan, United States

## Abstract

SARS-CoV-2 RNA in wastewater settled solids is associated with COVID-19 incidence in sewersheds and therefore, there is a strong interest in using these measurements to augment traditional disease surveillance methods. A wastewater surveillance program should provide rapid turn around for sample measurements (ideally within 24 hours), but storage of samples is necessary for a variety of reasons including biobanking. Here we investigate how storage of wastewater solids at 4°C, -20°C, and -80°C affects measured concentrations of SARS-CoV-2 RNA. We find that short term (7-8 d) storage of raw solids at 4°C has little effect on measured concentrations of SARS-CoV-2 RNA, whereas longer term storage at 4°C (35-122 d) or freezing reduces measurements by 60%, on average. We show that normalizing SARS-CoV-2 RNA concentrations by concentrations of pepper mild mottle virus (PMMoV) RNA, an endogenous wastewater virus, can correct for changes during storage as storage can have a similar effect on PMMoV RNA as on SARS-CoV-2 RNA. The reductions in SARS-CoV-2 RNA in solids during freeze thaws is less than those reported for the same target in liquid influent by several authors.

## Introduction

SARS-CoV-2 RNA in settled solids from wastewater treatment plants correlates to COVID-19 incidence in the sewershed population [1–3]. As a result, local and federal governmental agencies are establishing wastewater-based epidemiology methods to help inform pandemic response [4]. Wastewater consists of liquid and solid components. While many wastewater surveillance efforts have focused on measuring SARS-CoV-2 in the liquid component of wastewater [5,6], the solids have 10^3^ to 10^4^ higher concentrations of SARS-CoV-2 RNA on a per mass basis [2,7]. Settled solids are readily collected from the primary clarifier where they settle as part of the wastewater treatment process, or they can be settled from wastewater influent using standard method SM2540 F [8] if a wastewater treatment plant does not have a primary clarifier unit process.

In order for wastewater data on SARS-CoV-2 to be useful for real time disease response, samples should be analyzed quickly and results reported as soon as possible to public health officials. In such a scenario, samples are not stored, but are processed as soon as they are collected. Even if this is done as recommended, sample storage is essential. It may take 24 hours or longer for results from a sample to be obtained. In cases where an instrument malfunctions or results do not pass quality control metrics, samples might need to be rerun. Samples therefore need to be stored for at least as long as it takes to obtain results. Additionally, labs may want to create a biobank of samples; these samples can be used in the future to probe the presence of variants of concerns or other pathogens as needed.

A few studies have investigated how storage conditions affect quantification of SARS-CoV-2 RNA in liquid influent [6,9–12] and determined that storage and freeze thaws of the liquid influent can reduce measured concentrations of the viral RNA an order of magnitude or more. No published studies to date have investigated the effect of storage conditions on quantification of SARS-CoV-2 RNA in settled solids. Therefore, the goal of this study is to assess the impact of different realistic storage conditions on the quantification of SARS-CoV-2 RNA and an endogenous viral control (pepper mild mottle virus, PMMoV) in settled solids. The results of this study will inform optimal storage conditions for settled solids for use for wastewater-based epidemiology.

## Materials & Methods

### Sample collection

Eleven (11) 50-ml samples of settled solids were collected from the primary clarifiers at four unique wastewater treatment plants (Table 1) using sterile technique and clean containers. Samples were immediately stored on ice and transported to the lab where they were processed within 6 hours of sample pick up from the plants with high throughput methods [13– 15]. Thereafter, the samples were subjected to different storage treatments in the laboratory (Table 2). Raw samples were either stored at 4°C for 7-122 days, or 20°C for 2-3 d. Four samples were stored as dewatered solids (described below) at -80°C for 35 to 122 days. After storage, refrigerated samples were immediately processed, and frozen samples were removed from the freezers and defrosted at 4°C for 24 hours and then processed according to Topol et al. [13–15]. In total, there were 16 samples subjected to a storage treatment.

**Table 1.** Samples used in this study as well as the wastewater plant they were collected from (all located in California) and the date of sample collection, and the storage temperature and duration for the treatment applied to it.

| Sample ID | Plant | Date | Storage condition(s) | duration(s) of storage |
| --- | --- | --- | --- | --- |
| SJ200080 | SJ | 2/28/21 | 4°C | 8 d |
| D300230 | D | 3/1/21 | 4°C | 7 d |
| OSP700097 | Ocean | 3/1/21 | 4°C | 7 d |
| SJ200082 | SJ | 2/23/21 | -20°C | 3 d |
| D300234 | D | 2/24/21 | -20°C | 2 d |
| OSP700126 | Ocean | 2/24/21 | -20°C | 2 d |
| SAC900313 | SAC | 3/1/21 | 4°C / -20°C | 7 d / 2 d * |
| SJ200128 | SJ | 2/25/21 | 4°C / -80°C | 35 d |
| SJ20023 | SJ | 11/30/20 | 4°C / -80°C | 122 d |
| D300243 | D | 2/25/21 | 4°C / -80°C | 35 d |
| D300066 | D | 12/31/20 | 4°C / -80°C | 91 d |

\*The sample stored at -20°C was stored for 2 d and the sample stored at 4°C was stored for 7 d.

**Table 2.**
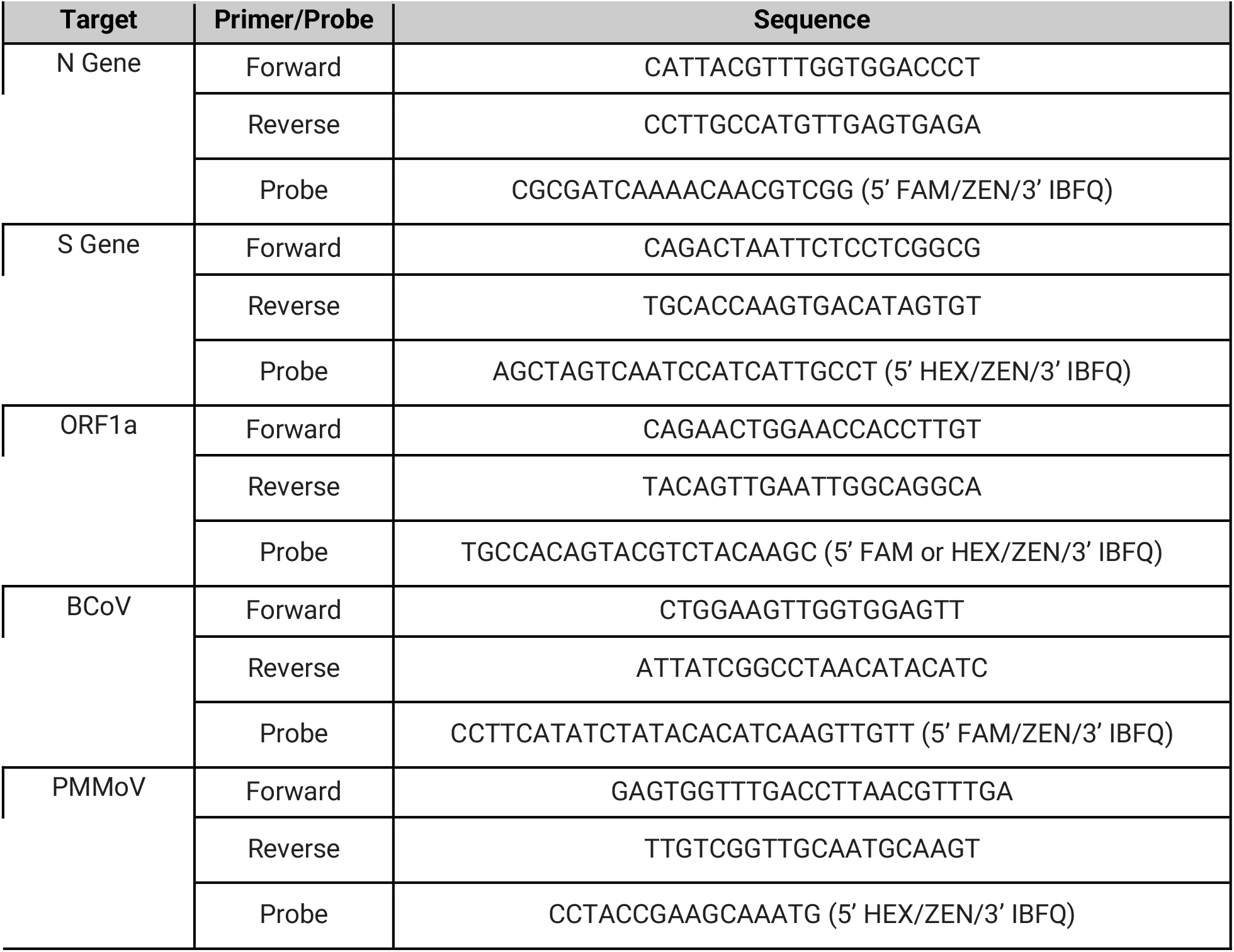
The molecular targets used in this study as well as the primer and probe sequences. The N, S, and ORF1a genes are located within SARS-CoV-2 genome. The BCoV target is for bovine coronavirus, a process control spiked into the sample during processing. PMMoV is for the internal endogenous control which is naturally present in high concentrations in the samples. Additional details of these assays can be found in Huisman et al. [22]

### Sample Preparation

The solids were dewatered by centrifugation at 24,000 x g for 30 minutes at 4°C. The supernatant was aspirated and discarded. A 0.5 - 1 g aliquot of the dewatered solids was dried at 110°C for 19-24 hrs to determine its dry weight. Bovine coronavirus (BCoV) was used as a positive recovery control. Each day, attenuated bovine coronavirus vaccine (PBS Animal Health, Calf-Guard Cattle Vaccine) was spiked into DNA/RNA shield solution (Zymo Research) at a concentration of 1.5 µL /mL. Dewatered solids were resuspended in the BCoV-spiked DNA/RNA shield to a concentration of 75 mg/mL. This concentration of solids was chosen as previous work titrated solutions with varying concentrations of solids to identify a concentration at which inhibition of the SARS-CoV-2 assays was minimized (data not shown). 5-10 5/32” Stainless Steel Grinding Balls (OPS Diagnostics) were added to each sample which was subsequently homogenized by shaking with a Geno/Grinder 2010 (Spex SamplePrep). Samples were then briefly centrifuged to remove air bubbles introduced during the homogenization process, and then vortexed to re-mix the sample.

### RNA Extraction

RNA was extracted from 10 replicate aliquots per sample. For each replicate, RNA was extracted from 300 µl of homogenized sample using the Chemagic™ Viral DNA/RNA 300 Kit H96 for the Perkin Elmer Chemagic 360 followed by PCR Inhibitor Removal with the Zymo OneStep-96 PCR Inhibitor Removal Kit. Extraction negative controls (water) and extraction positive controls (500 copies of SARS-CoV-2 genomic RNA (ATCC) in the BCoV-spiked DNA/RNA shield solution described above) were extracted using the same protocol as the homogenized samples.

### Digital PCR

RNA extracts were used as template in digital droplet RT-PCR assays for SARS-CoV-2 N, S, and ORF1a RNA gene targets in a triplex assay, and BCoV and PMMoV in a duplex assay (see Table 3 for primer and probe sequences, purchased from IDT). Undiluted extract was used for the SARS-CoV-2 assay template and a 1:100 dilution of the extract was used for the BCoV / PMMoV assay template. Digital RT-PCR was performed on 20 µl samples from a 22 µl reaction volume, prepared using 5.5 µl template, mixed with 5.5 µl of One-Step RT-ddPCR Advanced Kit for Probes (Bio-Rad 1863021), 2.2 µl Reverse Transcriptase, 1.1 µl DTT and primers and probes at a final concentration of 900nM and 250nM respectively. Droplets were generated using the AutoDG Automated Droplet Generator (Bio-Rad). PCR was performed using Mastercycler Pro with the following protocol: reverse transcription at 50°C for 60 minutes, enzyme activation at 95°C for 5 minutes, 40 cycles of denaturation at 95°C for 30 seconds and annealing and extension at either 59°C (for SARS-CoV-2 assay) or 56°C (for PMMoV/BCoV duplex assay) for 30 seconds, enzyme deactivation at 98°C for 10 minutes then an indefinite hold at 4°C. The ramp rate for temperature changes were set to 2°C /second and the final hold at 4°C was performed for a minimum of 30 minutes to allow the droplets to stabilize. Droplets were analyzed using the QX200 Droplet Reader (Bio-Rad). All liquid transfers were performed using the Agilent Bravo (Agilent Technologies).

**Table 3.**
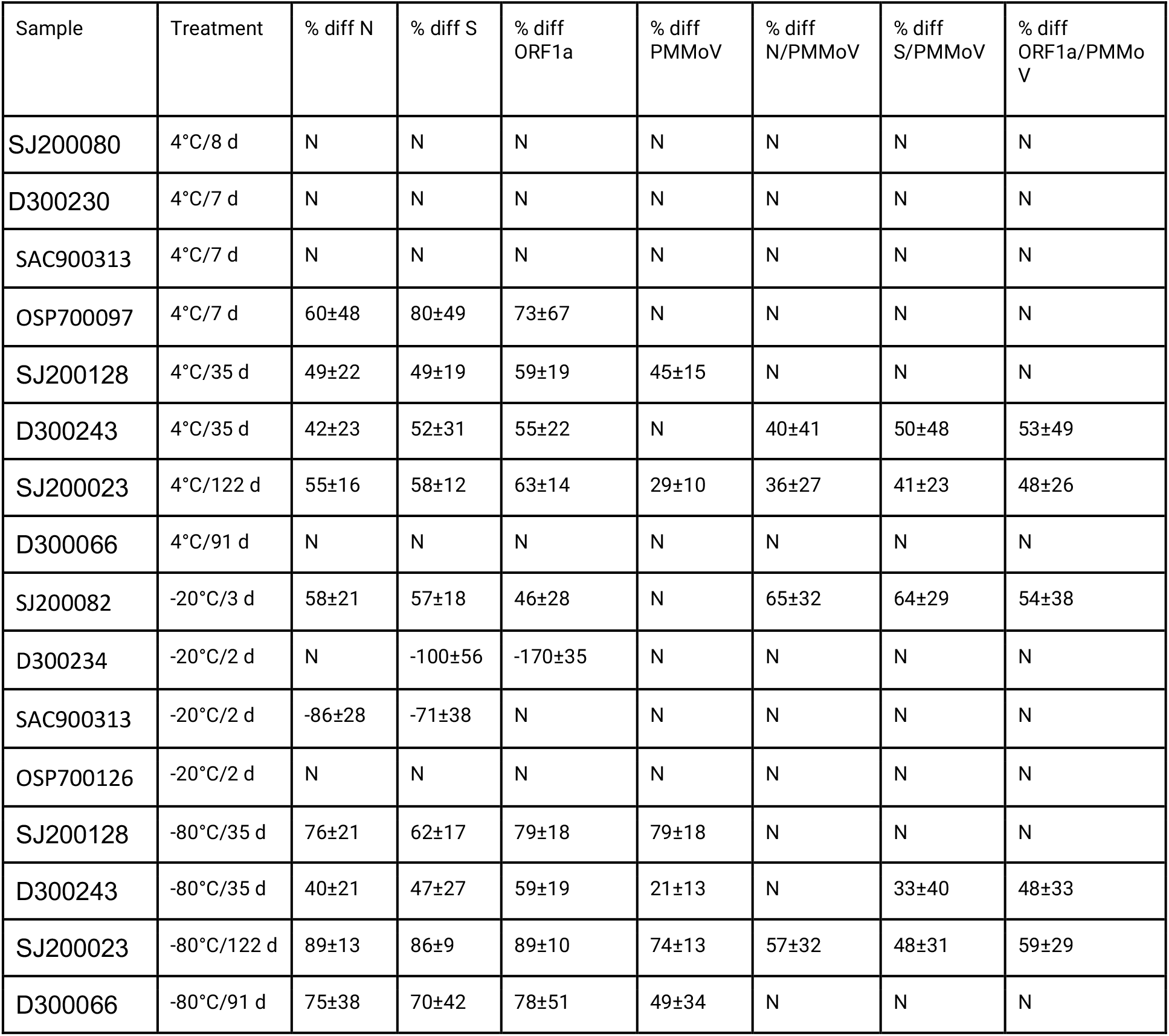
Results for comparisons between experimental treatments and controls.The percent difference (% diff) in SARS-CoV-2 RNA measured in treatments versus their control is shown then the difference was significantly different. “N” indicates that measurements or ratios were not different. The value after the ± is the standard deviation propagated from the measurements used to make the calculation. A positive percent difference indicates the treatment was lower than the control, a negative percent difference indicates the treatment was higher in the control. See methods for more details on the calculations.

| Sample | Treatment | % diff N | % diff S | % diff ORF1a | % diff PMMoV | % diff N/PMMoV | % diff S/PMMoV | % diff ORF1a/PMMoV |
| --- | --- | --- | --- | --- | --- | --- | --- | --- |
| SJ200080 | 4°C/8 d | N | N | N | N | N | N | N |
| D300230 | 4°C/7 d | N | N | N | N | N | N | N |
| SAC900313 | 4°C/7 d | N | N | N | N | N | N | N |
| OSP700097 | 4°C/7 d | 60±48 | 80±49 | 73±67 | N | N | N | N |
| SJ200128 | 4°C/35 d | 49±22 | 49±19 | 59±19 | 45±15 | N | N | N |
| D300243 | 4°C/35 d | 42±23 | 52±31 | 55±22 | N | 40±41 | 50±48 | 53±49 |
| SJ200023 | 4°C/122 d | 55±16 | 58±12 | 63±14 | 29±10 | 36±27 | 41±23 | 48±26 |
| D300066 | 4°C/91 d | N | N | N | N | N | N | N |
| SJ200082 | -20°C/3 d | 58±21 | 57±18 | 46±28 | N | 65±32 | 64±29 | 54±38 |
| D300234 | -20°C/2 d | N | -100±56 | -170±35 | N | N | N | N |
| SAC900313 | -20°C/2 d | -86±28 | -71±38 | N | N | N | N | N |
| OSP700126 | -20°C/2 d | N | N | N | N | N | N | N |
| SJ200128 | -80°C/35 d | 76±21 | 62±17 | 79±18 | 79±18 | N | N | N |
| D300243 | -80°C/35 d | 40±21 | 47±27 | 59±19 | 21±13 | N | 33±40 | 48±33 |
| SJ200023 | -80°C/122 d | 89±13 | 86±9 | 89±10 | 74±13 | 57±32 | 48±31 | 59±29 |
| D300066 | -80°C/91 d | 75±38 | 70±42 | 78±51 | 49±34 | N | N | N |

Each sample was run in 10 replicate wells, extraction negative controls were run in 7 wells, and extraction positive controls in 1 well. In addition, PCR positive controls for SARS-CoV-2 RNA, BCoV, and PMMoV were run in 1 well, and NTC were run in 7 wells. Positive controls consisted of BCoV and PMMoV gene block controls (purchased from IDT) and gRNA of SARS-CoV-2 (ATCC^®^ VR-1986D™). Results from replicate wells were merged for analysis. Thresholding was done using QuantaSoft™ Analysis Pro Software (Bio-Rad, version 1.0.596). Additional details are provided in supporting material per the dMIQE guidelines [16].

### Data analysis

Concentrations of RNA targets were converted to concentrations per dry weight of solids in units of copies/g dry weight. The total error is reported as 68% confidence intervals and includes the errors associated with the poisson distribution and the variability among the 10 replicates. The recovery of BCoV was determined by normalizing the concentration of BCoV by the expected concentration given the value measured in the spiked DNA/RNA shield. If the BCoV recovery was less than 10%, then the sample was rerun.

PMMoV, N, S, ORF1a, as well as N/PMMoV, S/PMMoV, and ORF1a/PMMoV were compared for each sample control and treatment, 1 by 1, by examining the measurement and the 68% error associated with the measurement. The error associated with the quotients was estimated by propagating errors on the numerator and denominator. If the measurement of the treatment condition fell within the error range of the control condition, then the measurement was deemed “not different”. This approach is equivalent to a t-test where the null hypothesis (Ho) is the value of the treatment is the same as the control and the alternate hypothesis (Ha) is that the values are different. In this study, we are particularly concerned about type 2 errors (failing to reject Ho when it is false) as we are concerned with whether storage renders different measurements. As such, in order to increase the power of the analysis, we chose to make comparisons using the 68% confidence intervals. With the 10 replicates, this gives ∼90% power of avoiding a type 2 error assuming an effect size equal to the standard deviation.

For measurements deemed “different”, the percent difference (% diff) was calculated as % diff = 100 x (control-treatment)/control where control and treatment are the associated measurements. A positive % diff indicates that the treatment result is smaller than the control, whereas a negative percent indicates the treatment had higher concentrations than the control. Errors for % diff were propagated from the measurements as standard deviations.

## Results

### Quality control

All positive extraction and PCR controls were positive, and all negative extraction controls and negative PCR controls were negative indicating no cross contamination between samples. Recovery of spiked BCoV was above 10% for all samples so no samples were rerun owing to unacceptable recovery. SARS-CoV-2 RNA and PMMoV targets were detected in all samples. Data from the experiments is available through Stanford Digital Repository [17].

### Short-term Storage at 4°C

Two experiments were carried out to investigate the effects of storage on 8 samples of raw settled solids samples at 4°C. First, all samples were processed within 6 hours of collection and the resultant measurements were treated as those of the control condition. Then, in one experiment, 4 samples were stored for 7-8 days at 4°C prior to being processed a second time; and in the other, 4 samples were stored between 35 days and 122 days prior to being processed a second time. The stored conditions are referred to as treatments.

SARS-CoV-2 RNA measurements made after 7-8 d of storage at 4°C were not different from the control condition for 3 of the 4 samples; PMMoV was not different between treatment and control for all 4 of the samples. When SARS-CoV-2 RNA gene concentrations were normalized by PMMoV gene concentrations, the ratios were not different between treatment and control in any of the 4 samples (Figure 1). For the sample that had lower SARS-CoV-2 RNA gene concentrations in the treatment compared to the control, the concentrations of the N, S, and ORF1 genes differed by 60%, 80%, and 73% respectively (Table 3).

**Figure 1.**
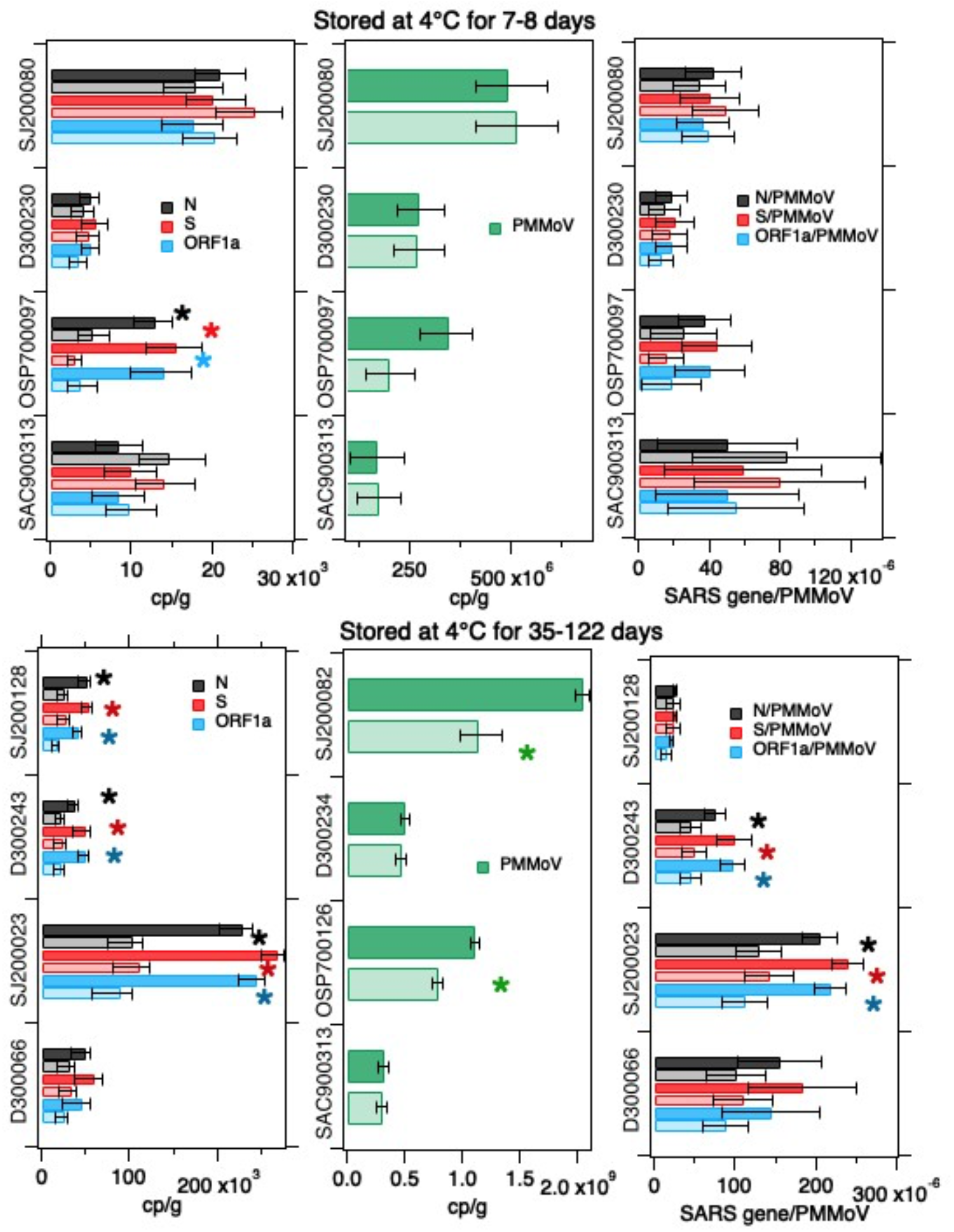
Concentrations of SARS-CoV-2 RNA targets (N, S, ORF1a) and PMMoV RNA, as well as their ratios as measured in settled solids in controls (darker bars) and treatments (lighter bars) and total errors as reported by the digital PCR instrument, or in the case of the ratios, the errors were propagated. Asterisks denote measurements for which the errors on the treatments and controls do not overlap and indicate the measurements were different. Top row shows experiments where solids were stored at 4°C for 7-8 days while the bottom row shows experiments where solids were stored at 4°C for 35-122 days (see Table 1).

### Long-term Storage at 4°C

Longer storage at 4°C (between 35 and 122 days) resulted in significantly lower measurements of SARS-CoV-2 RNA in 3 of the 4 tested samples, and lower PMMoV RNA concentrations in 2 of 4 samples. Normalizing SARS-CoV-2 RNA concentrations by PMMoV concentrations “corrected” the differences observed in 1 sample, but not the other 2. For those 2 samples, ratios were lower and different in the treatments compared to the controls (Figure 1). The difference between the treatment and controls are shown in Table 3. Generally, the differences in the measurements, when they were different, were ∼50%.

### Storage at -20°C

Four samples were processed within 6 hours of collection to obtain control measurements. The same samples were also frozen at -20°C for 2 or 3 days, and then defrosted and processed to obtain treatment measurement.

SARS-CoV-2 RNA concentrations in treatments were different in 3 of the 4 samples compared to controls (lower in treatment versus control in 1 and higher in 2). PMMoV concentrations were not different between treatments and controls. Normalizing SARS-CoV-2 RNA concentrations by PMMoV RNA concentrations, “corrected” the differences in SARS-CoV-2 RNA observed in 2 of the 3 samples (Figure 2). Differences between the experiments are shown in Table 4; differences were between ∼60% and 100%.

**Figure 2.**
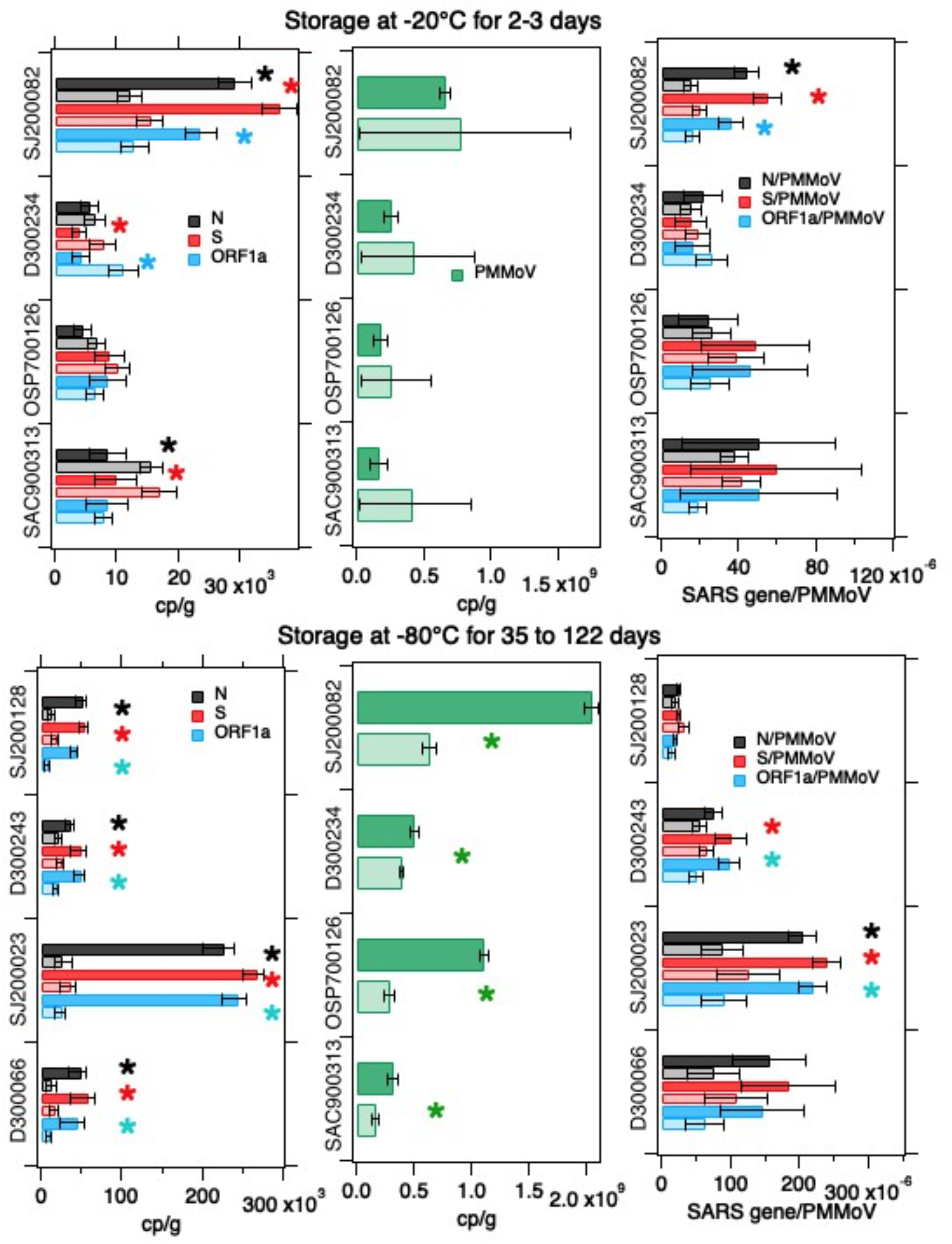
Concentrations of SARS-CoV-2 RNA targets (N, S, ORF1a) and PMMoV RNA, as well as their ratios as measured in settled solids in controls (darker bars) and treatments (lighter bars) and total errors as reported by the digital PCR instrument, or in the case of the ratios, the errors were propagated. Asterisks denote measurements for which the errors on the treatments and controls do not overlap and indicate the measurements were different. Top row shows experiments where solids were stored at -20°C for 2-3 days while the bottom row shows experiments where dewatered solids were stored at -80°C for 35-122 days (see Table 1).

### Storage at -80°C

Four samples were processed within 6 hours of collection to obtain control measurements and also dewatered,, frozen at -80°C for between 35 and 122 days, and then defrosted and processed to obtain treatment measurements.

SARS-CoV-2 and PMMoV RNA concentrations were statistically lower in the treatments compared to the controls by 40% to 90%. Normalizing SARS-CoV-2 RNA concentrations by PMMoV concentrations “corrected” the differences for 2 of the 4 samples. For the other 2, the ratio of SARS-CoV-2 RNA to PMMoV RNA concentrations was ∼50% lower in the treatment than the control.

## Discussion

SARS-CoV-2 RNA in settled solids correlates to COVID-19 incidence in surrounding sewersheds [1–3]. While immediate processing of samples is ideal, it is necessary to store and archive samples in case samples need to be rerun due to failed quality control metrics, or if additional analyses are needed to investigate the presence of other viruses or viral variants, for example. Our experiments suggest that storage of samples, either refrigerated or frozen, may change the concentrations of SARS-CoV-2 RNA measured in the samples, but changes are less than an order of magnitude. Across the treatment/samples where we observed a change relative to the control (n=11 of 16), there was a reduction in measured SARS-CoV-2 RNA concentration in all but 2 sample treatments (mean percent reduction of 63% ±14% standard deviation, n = 27: 9 treatment/samples x 3 SARS-CoV-2 gene measurements per treatment/sample). In the 2 cases where we observed an increase in SARS-CoV-2 concentrations in the treatment compared to the control, that increase was 107%±44% on average (n = 4, 2 sample /treatments x 2 SARS-CoV-2 genes were different per sample/treatment). All measurements made in samples receiving a treatment were within an order of magnitude of the measurement in the control.

Several studies have examined the effect of sample storage on SARS-CoV-2 quantification in wastewater influent which represents a different matrix than that examined herein. Influent consists primarily of liquid wastewater rather than solids. Hokajärvi et al. [10] examined the effect of storage on detection of SARS-CoV-2 RNA in liquid influent and found small differences resulting from storage at freezing temperatures and first order decay of the RNA in influent stored at 4°C with a T_90_ (time until 1 log_10_ reduction of concentration) of 36 to 52 days depending on the genomic target. Ahmed et al. [12] report similar results as Hokajärvi et al. for decay of the SARS-CoV-2 targets in influent during storage at 4°C. Markt et al. [18] found minimal differences in SARS-CoV-2 RNA concentrations measured in influent stored for up to 7 days at 4°C compared to concentrations measured with no storage, but found more than an order of magnitude decrease in SARS-CoV-2 RNA concentrations in samples that were stored frozen and subject to a freeze thaw. Fernandez-Cassi et al. [19] report extensive reduction in SARS-CoV-2 RNA concentrations measured in liquid wastewater stored at 4°C and -20°C (1-2 orders of magnitude). Our results regarding solids stored at 4°C are similar to those presented in these influent studies, except for Fernandez-Cassi et al. [19]; overall we saw minimal reduction (less than an order of magnitude) even for samples stored over 100 days. However, the effect of freeze thaw on our measurements with solids is small compared to those reported by Markt et al. [18] and Fernandez-Cassi et al. [19] for infuent. We could identify only one published study on SARS-CoV-2 RNA decay in solids: Hokajärvi et al. [10] report minimal decay of SARS-CoV-2 RNA in a “pellet” consisting of settled solids from influent during storage at 4°C, -20°C, and -75°C.

Researchers have used PMMoV as an internal process control in their efforts to monitor SARS-CoV-2 in wastewater [1,20,21]. Assuming endogenous PMMoV RNA is recovered in the sample processing and RNA extraction and purification process in the same manner as SARS-CoV-2 RNA, then normalizing SARS-CoV-2 RNA by PMMoV RNA provides a ratio that does not depend on recovery. Wolfe et al. [1] showed the ratio of SARS-CoV-2 RNA/PMMoV RNA in settled solids is associated with COVID-19 incidence rates empirically, and the relationship between the ratio and COVID-19 incidence rates also falls from a mass balance model that relates wastewater solid concentrations to the number of people shedding SARS-CoV-2 RNA in their stool.

In this study, we found that normalizing SARS-CoV-2 RNA by PMMoV corrected for changes in concentration that may result during storage. In 7 of the 11 sample/treatments that showed differences between SARS-CoV-2 RNA concentrations and the control, SARS-CoV-2/PMMoV was not different between treatment and control. When there were differences, they were less than an order of magnitude. PMMoV RNA concentration was often affected by storage in a similar manner as SARS-CoV-2 RNA concentrations, thus highlighting an additional benefit of using the internal control to interpret concentrations of the SARS-CoV-2 RNA targets in wastewater-based epidemiology applications. The ability to effectively correct for the impact of storage on samples suggests that the primary concerns for sample storage of wastewater solids are related to times when SARS-CoV-2 concentrations are nearing the limit of detection, and during periods of low incidence immediate sample processing should be a higher priority.

Additional research should examine a time course of decay for a single solids sample stored for various lengths of time, and also investigate the effects of multiple freeze thaws on target quantification. Work with additional viral targets may also be useful to provide guidance on storage for wastewater-based epidemiology applications beyond COVID-19. Finally, our study was not powered to investigate whether storage affected viral quantification in wastewater solids from different wastewater treatment plants in different ways; we used samples from diverse plants to capture potential variations between properties of wastewater solids. Additional work should investigate if storage has differential effects on samples from different wastewater treatment plants.

## Conclusions

Storage of wastewater solids for use in wastewater-based epidemiology applications is essential. It is therefore important to understand how storage affects concentrations of health-relevant targets. Here we examined how storage at 4°C for short (7-8 d) and long durations (35-122 d), -20°C for short (2-3 d), and -80°C for long durations (35-122 d) affects SARS-CoV-2 RNA measurements in wastewater solids, and whether normalizing measurements by concentrations of an internal control corrects for the effects of storage.

We found storage at 4°C for short durations of 7-8 days had limited to no effect on measured concentrations, but other storage conditions and durations affected concentrations by reducing them by ∼60%, on average, and in one case increasing them by up to 170%. However, we found that the normalizing concentrations by the internal control PMMoV corrected for the observed differences in many cases.

As such, we recommend short duration storage at 4°C, and normalizing concentrations of SARS-CoV-2 RNA by concentrations of PMMoV in the sample. Even under the longer storage conditions including those that required a freeze/thaw, changes in concentrations observed with the solids were less than one order of magnitude and similar among samples subjected to the same treatment.

## Data Availability

Data are available from Stanford Digital Repository at the link provided in the paper.

https://purl.stanford.edu/yn042kx5009

## Acknowledgements

We acknowledge the wastewater treatment plant staff for providing the samples. This study was performed on the ancestral and unceded lands of the Muwekma Ohlone people. We pay our respects to them and their Elders, past and present, and are grateful for the opportunity to live and work here.

