## supporting material for "Effect of storage conditions on SARS-CoV-2 RNA quantification in wastewater solids"

The average (standard deviation) number of droplets in ten merged wells determined from a random subset of 8 samples was 190000 (6800). Average number of copies per partition (λ) for SARS-CoV-2 RNA in the same subset was $7.2 \times 10^{-4}$, the average of the number of copies per partition (λ) for PMMoV was 0.14, and $1.8 \times 10^{-3}$ for BCoV.

As the samples were extracted ten times and each extract analyzed in one of 10 replicate wells which were merged, the replicate variability incorporates variation from both RNA extraction and RT-dPCR with a heterogeneous solids sample. Sample standard deviations for the SARS-CoV-2, PMMoV RNA, and BCoV RNA quantification estimated from the merged wells were, on average 21%, 17%, and 15% of the measurement. Assays were conducted in only one lab, so reproducibility was not assessed.

The LOD of the assay was 3 positive droplets which translates into between 600 and 1000 copies/g dry weight depending on the percent dry weight of the solids used in the extraction which depends on the properties of the solid and how effectively it can be dewatered.

Example fluorescence plots for the N, S, and ORF1a genes are provided in the associated reference by Topol et al. [1], additional fluorescence plots are shown below in Figures S1, S2, and S3.

| ITEM TO CHECK | PROVIDED | COMMENT |
|---|---|---|
| | Y/N | |
| 1. SPECIMEN | | |
| Detailed description of specimen type and numbers | Y | Methods of Main Text |
| Sampling procedure (including time to storage) | Y | Methods of Main Text |
| Sample aliquotation, storage conditions and duration | Y | Methods of Main Text |
| 2. NUCLEIC ACID EXTRACTION | | |
| Description of extraction method including amount of sample processed | Y | Methods of Main Text |
| Volume of solvent used to elute/resuspend extract | Y | Methods of Main Text |
| Number of extraction replicates | Y | Methods of Main Text |
| Extraction blanks included? | Y | Methods of Main Text |
| 3. NUCLEIC ACID ASSESSMENT AND STORAGE | | |
| Method to evaluate quality of nucleic acids | N | Not Done |
| Method to evaluate quantity of nucleic acids (including molecular weight and | N | Not Done |
| Storage conditions: temperature, concentration, duration, buffer, aliquots | Y | Methods of Main Text |
| Clear description of dilution steps used to prepare working DNA solution | Y | Methods of Main Text |
| 4. NUCLEIC ACID MODIFICATION | | |
| Template modification (digestion, sonication, pre-amplification, bisulphite | N | NA |
| Details of repurification following modification if performed | Y | Zymo Column in Methods of Main Text |
| 5. REVERSE TRANSCRIPTION | | |
| cDNA priming method and concentration | N | NA |
| One or two step protocol (include reaction details for two step) | Y | Methods of Main Text |
| Amount of RNA added per reaction | Y | Methods of Main Text |
| Detailed reaction components and conditions | Y | Methods of Main Text, also provided in protocols.io protocol referenced in main text |
| Estimated copies measured with and without addition of RT* | N | Not Done |
| Manufacturer of reagents used with catalogue and lot numbers | N | Reagents, Manufacturers, and Catalogue Numbers Reported in Supplemental Material. Lot Numbers are Not Reported |
| Storage of cDNA: temperature, concentration, duration, buffer and aliquots | Y | NA |
| 6. dPCR OLIGONUCLEOTIDES DESIGN AND TARGET INFORMATION | | |
| Sequence accession number or official gene symbol | Y | Provided in reference in paper to Huisman et al. |
| Method (software) used for design and *in silico* verification | Y | Provided in reference in paper to Huisman et al. |
| Location of amplicon | Y | Provided in reference in paper to Huisman et al. |
| Amplicon length | Y | Provided in reference in paper to Huisman et al. |
| Primer and probe sequences (or amplicon context sequence)** | Y | Methods in Main Text |
| Location and identity of any modifications | Y | NA |
| Manufacturer of oligonucleotides | Y | Methods in Main Text |
| 7. dPCR PROTOCOL | | |
| Manufacturer of dPCR instrument and instrument model | Y | Methods of Main Text, also provided in protocols.io protocol |
| Buffer/kit manufacturer with catalogue and lot number | Y | Methods of Main Text, also provided in protocols.io protocol |
| Primer and probe concentration | Y | Methods of Main Text, also provided in protocols.io protocol |
| Pre-reaction volume and composition (incl. amount of template and if | Y | Methods of Main Text, also provided in protocols.io protocol |
| Template treatment (initial heating or chemical denaturation) | N | NA |
| Polymerase identity and concentration, Mg++ and dNTP concentrations*** | N | Included in Kit Manuals |
| Complete thermocycling parameters | Y | Methods of Main Text, also provided in protocols.io protocol referenced in main text |
| 8. ASSAY VALIDATION | | |
| Details of optimisation performed | N | Commercial Kit, Followed Manufacturer's Instructions |
| Analytical specificity (vs. related sequences) and limit of blank (LOB) | N | Provided in reference in paper to Huisman et al. |
| Analytical sensitivity/LoD and how this was evaluated | Y | Methods of Main Text |
| Testing for inhibitors (from biological matrix/extraction) | Y | Methods of Main Text |
| 9. DATA ANALYSIS | | |
| Description of dPCR experimental design | Y | Methods of Main Text |
| Comprehensive details negative and positive of controls (whether applied for | Y | Methods of Main Text |
| Partition classification method (thresholding) | Y | Methods of Main Text, also provided in protocols.io protocol referenced in main text |
| Examples of positive and negative experimental results (including | Y | Supplemental Information |
| Description of technical replication | Y | Methods of Main Text |
| Repeatability (intra-experiment variation) | Y | Supplemental Information |
| Reproducibility (inter-experiment/user/lab etc. variation ) | N | Assays were only completed within one laboratory |
| Number of partitions measured (average and standard deviation ) | Y | Supplemental Information |
| Partition volume | Y | Reported by Manufacturer |
| Copies per partition (λ or equivalent ) (average and standard deviation) | Y | Supplemental Information |
| dPCR analysis program (source, version) | Y | Methods of Main Text |
| Description of normalisation method | Y | Methods of Main text when applicable |
| Statistical methods used for analysis | Y | Methods of Main Text |
| Data transparency | raw data uploaded to online repository with ID: | https://purl.stanford.edu/yn042kx5009 |

20

Table S1. dMIQE2020 checklist for authors, reviewers and editors. Authors should fill detail whether information is provided. Where ‘yes’ is selected use comment box to detail location of information or to include the information. Where ‘no’ is selected use comment box to outline rationale for omission. Sections 4 and 5 may not apply depending on experiment. From Huggett et al. [2]. Huisman et al. [3] is cited in the table.

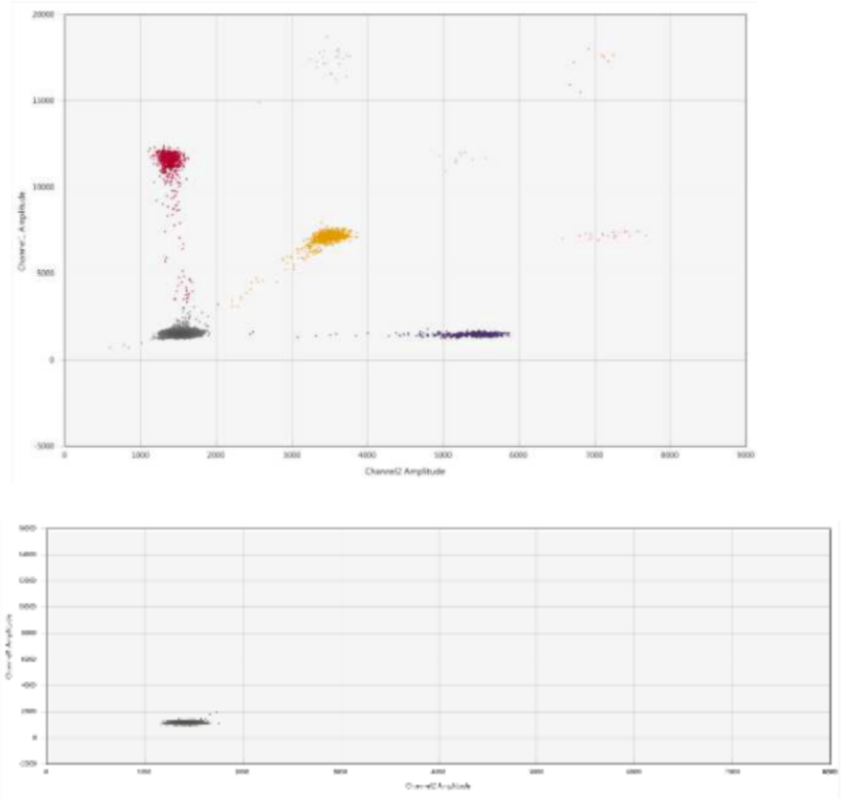


**Fig. S1: Fluorescence plots from the Bio-Rad QX200 used for the wastewater solids samples for N, S, and ORF1a.** Positive experimental results are provided in the top image and negative experimental results are shown in the bottom image.

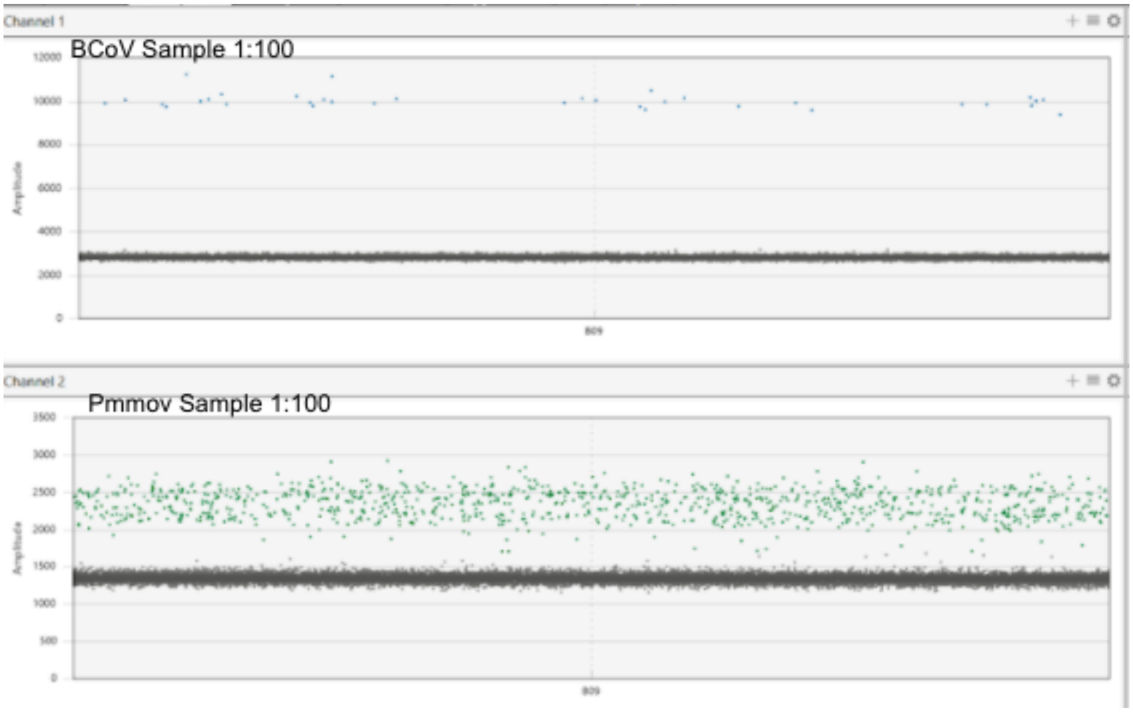


**Fig. S2: Fluorescence plots from the Bio-Rad QX200 used for the wastewater solids samples for PMMoV and BCov.** Positive experimental results are shown.

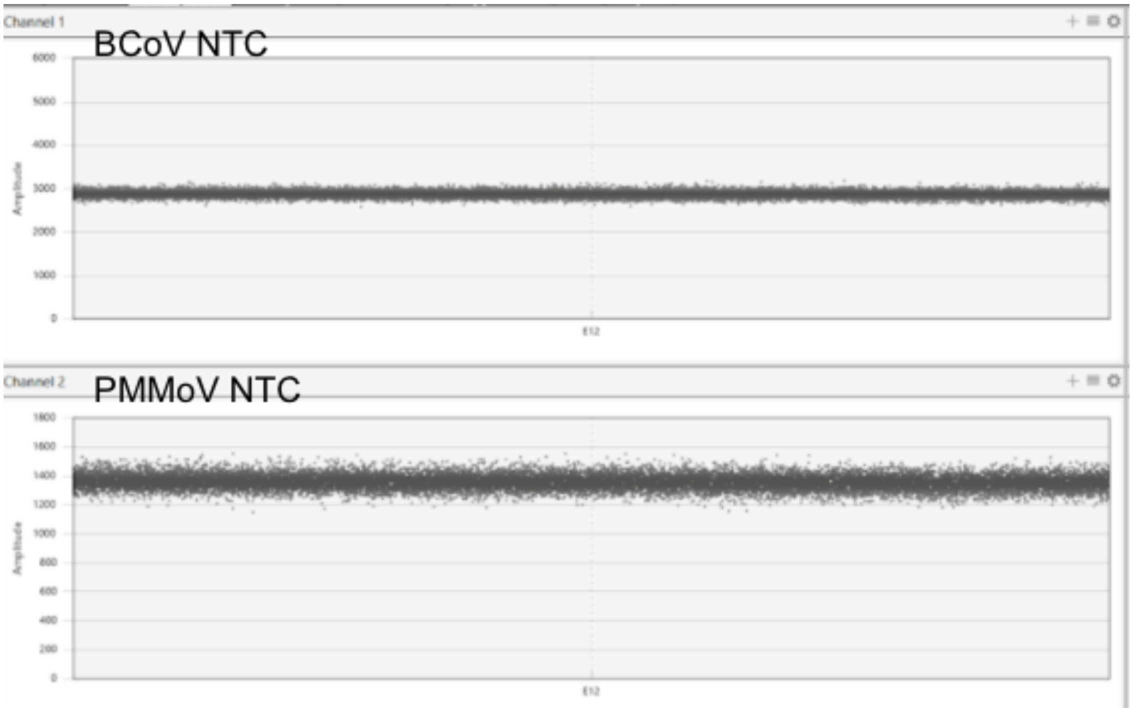


**Fig. S3: Fluorescence plots from the Bio-Rad QX200 used for the wastewater solids samples for PMMoV and BCov.** Negative experimental results are shown.
